# ARE LLMS READY FOR PEDIATRICS? A COMPARATIVE EVALUATION OF MODEL ACCURACY ACROSS CLINICAL DOMAINS

**DOI:** 10.1101/2025.04.25.25326437

**Authors:** Gianluca Mondillo, Simone Colosimo, Alessandra Perrotta, Vittoria Frattolillo, Mariapia Masino

## Abstract

Large Language Models (LLMs) are rapidly emerging as promising tools in the healthcare field, yet their effectiveness in pediatric contexts remains underexplored. This study evaluated the performance of eight contemporary LLMs, released between 2024 and 2025, in answering multiple-choice questions from the MedQA dataset, stratified into two distinct categories: adult medicine (1461 questions) and pediatrics (1653 questions). Models were tested using a standardized prompting methodology with default hyperparameters, simulating real-world use by non-expert clinical users. Accuracy scores for adult and pediatric subsets were statistically compared using the chi-square test with Yates’ correction. Five models (Amazon Nova Pro 1.0, GPT 3.5-turbo-0125, Gemini 2.0 Flash, Grok 2, and Claude 3 Sonnet) demonstrated significantly lower performance on pediatric questions, with accuracy drops of up to more than 10 percentage points. In contrast, ChatGPT-4o, Gemini 1.5 Pro, and Claude 3.5 Sonnet showed comparable performance across both domains, with ChatGPT-4o achieving the most balanced result (accuracy: 83.57% adult, 83.18% pediatric; p = 0.80). These findings suggest that while some models struggle with pediatric-specific content, more recent and advanced LLMs may offer improved generalizability and domain robustness. The observed variability highlights the critical importance of domain-specific validation prior to clinical implementation, particularly in specialized fields such as pediatrics.

## 1 Introduction

Large Language Models (LLMs) represent a rapidly evolving technological frontier, with growing interest in their potential applications within the healthcare sector. Their ability to understand and generate complex natural language positions them as promising tools for decision support, continuing medical education, and improved access to medical information for both professionals and patients. Despite their promise, the safe and effective integration of these tools requires rigorous validation of their performance [1], especially in specialized clinical fields such as pediatrics. This area stands apart from adult medicine due to a range of critical factors: age-specific disease etiologies, dynamic age-dependent physiological parameters and laboratory reference ranges, weight- and body surface area-based drug dosing principles, and unique considerations related to physical and cognitive development.

Our working hypothesis is that earlier-generation LLMs, or those built on potentially less sophisticated architectures—such as models with fewer parameters or less advanced pre-training techniques—might exhibit lower performance when tackling pediatric-specific questions compared to those focused on adult medicine. This discrepancy could stem from training corpora that predominantly reflect adult medicine or from an intrinsic limitation in the model’s ability to abstract and apply knowledge to pediatric clinical contexts that are underrepresented in general training data. Pediatric-specific dosages, rare syndromes, and the differing clinical presentations of common diseases in children could constitute particular points of weakness.

This study aims to conduct a quantitative comparative analysis of the capabilities of a diverse panel of contemporary LLMs in responding to multiple-choice questions (MCQs) drawn from the MedQA dataset [2, 3], carefully stratified into adult medicine and pediatrics domains. The analysis seeks to determine the existence and statistical significance of any performance discrepancies between the two domains for each model tested and to explore possible correlations between these discrepancies and the known or presumed characteristics of the models.

## 2 Materials and Methods

### 2.1 Dataset

The MedQA dataset was used, a well-established resource containing multiple-choice questions modeled after medical licensing exams. For this study, only text-based questions were selected—those without graphical elements or images and featuring a single correct answer option.

### 2.2 Question Stratification

The selected questions were meticulously divided into two mutually exclusive subsets: “Adult Medicine” (Adult) and “Pediatrics” (Pediatric). Classification was performed by analyzing the prevailing clinical content of each question, identified either through the patient’s age explicitly mentioned in the question or implicitly inferred from the clinical context. The “Adult” set included 1,461 questions, while the “Pediatric” set contained 1,653.

### 2.3 Evaluated Models

The study included a representative sample of LLMs available at the time of analysis, selected to cover a range of architectures, developers, and release periods (Table 1 [4]).

**Table 1:** Comparison of major AI models released between 2024 and 2025, including release date, context window, architecture, developer, and knowledge cut-off.

| Model | Release Date | Context Window (input/output) | Architecture | Company | Knowledge Cut-off |
| --- | --- | --- | --- | --- | --- |
| Amazon Nova Pro 1.0 [5] | January 2025 | 300,000 / 50,000 tokens | Multimodal, optimized for efficiency | Amazon | Not specified |
| GPT-3.5-turbo-0125 [6] | 2024 | 16,385 / 4,096 tokens | Transformer | OpenAI | September 2021 |
| ChatGPT-4o [7] | May 2024 | 128,000 / 4,096 tokens | Multimodal | OpenAI | October 2023 |
| Gemini 1.5 Pro [8] | Feb 2024 | 1,000,000 / 32,000 tokens | Mixture-of-Experts (MoE) | Google | September 2024 |
| Gemini 2.0 Flash [9] | Dec 11, 2024 | 1,000,000 / 32,000 tokens | Multimodal, latency-optimized | Google | August 2024 |
| Grok 2 [10] | Aug 13, 2024 | 128,000 / 128,000 tokens | Not specified | xAI | Not specified |
| Claude 3 Sonnet (2024-02-29) [11] | Feb 29, 2024 | 200,000 / 4,096 tokens | Multimodal | Anthropic | August 2023 |
| Claude 3.5 Sonnet (2024-10-22) [12] | Oct 22, 2024 | 200,000 / 4,096 tokens | Multimodal | Anthropic | October 2024 |

### 2.4 Evaluation Procedure

Each LLM was tested on the entire set of stratified questions. The questions were presented to the models in batches of 25 at a time. A standardized and consistent prompting methodology was adopted for all models, instructing them to return only the letter corresponding to the answer option they deemed correct for each numbered question (e.g., ‘1. A’, ‘2. C’, …, ‘25. B’).

Access to the models was carried out through various interfaces, keeping hyperparameters unchanged to simulate standard usage by a non-expert user.

For ChatGPT-4o, the official web interface was used [13], with no possibility of modifying the system’s default hyperparameters, which are officially undisclosed. Gemini 1.5 Pro and Gemini 2.0 Flash were accessed through Google AI Studio [14], maintaining the platform’s default hyperparameters: Temperature = 1, Top P = 0.95, Max Output Tokens = 8192. For all other models (Amazon Nova Pro 1.0, GPT-3.5-turbo-0125, Grok 2, Claude 3 Sonnet, Claude 3.5 Sonnet), access was via the Chatbot Arena platform [15], using the interface’s default hyperparameters: Temperature = 0.7, Top P = 1, Max Output Tokens = 2048.

The response generated by each model (i.e., the chosen letter) was then programmatically compared to the official answer key of the MedQA dataset to determine correctness and calculate accuracy.

### 2.5 Statistical Analysis

For each LLM, accuracy was calculated separately for the “Adult” and “Pediatric” question sets as the ratio between the number of correct answers and the total number of questions in that category (Accuracy = Number of Correct Answers / Total Number of Questions). The statistical significance of the difference in proportions of correct answers between the two domains was assessed for each model using the Chi-square (*χ*^2^) test with Yates’ correction for continuity, comparing 2×2 contingency tables (Correct/Incorrect vs. Adult/Pediatric). A p-value below 0.05 was considered indicative of a statistically significant difference.

## 3 Results

The aggregated and comparative performances of the LLMs on the two question sets are detailed in Table 2.

**Table 2:** LLMs Performance on MedQA Adult vs Pediatric Questions.

| Model | Correct Answers (Adult) | Correct Answers (Pediatric) | Accuracy Adult | Accuracy Pediatric | Chi-square ($\chi^2$) | p-value | Significant Difference (p<0.05) |
| --- | --- | --- | --- | --- | --- | --- | --- |
| Amazon Nova Pro 1.0 | 1128/1461 | 1145/1653 | 77.20% | 69.26% | 24.39 | <0.00001 | YES |
| GPT 3.5-turbo-0125 | 943/1461 | 914/1653 | 64.54% | 55.59% | 27.19 | <0.00001 | YES |
| ChatGPT4o | 1221/1461 | 1375/1653 | 83.57% | 83.18% | 0.0596 | 0.80 | NO |
| Gemini 1.5 Pro | 1115/1461 | 1213/1653 | 76.31% | 73.38% | 3.14 | 0.07 | NO |
| Gemini 2.0 Flash | 1187/1461 | 1173/1653 | 81.24% | 70.96% | 44.13 | <0.00001 | YES |
| Grok 2 (2024-08-13) | 1303/1461 | 1431/1653 | 89.18% | 86.57% | 4.71 | 0.02 | YES |
| Claude 3 Sonnet (2024-02-29) | 951/1461 | 941/1653 | 65.09% | 56.90% | 21.34 | <0.0001 | YES |
| Claude 3.5 Sonnet (2024-10-22) | 1235/1461 | 1355/1653 | 84.50% | 81.90% | 3.44 | 0.06 | NO |

Figure 1 provides a graphical representation of accuracy per model. A notable heterogeneity in overall performance is observed across the evaluated models. Grok 2 emerges as the best-performing model overall on both datasets (Adult Accuracy: 89.18%; Pediatric Accuracy: 86.57%). At the other end of the spectrum, GPT 3.5-turbo-0125 shows the lowest accuracy scores (Adult: 64.54%; Pediatric: 55.59%). Statistical analysis reveals that for five of the eight tested models—specifically Amazon Nova Pro 1.0, GPT 3.5-turbo-0125, Gemini 2.0 Flash, Grok 2, and Claude 3 Sonnet (version 20240229)—the accuracy difference between adult and pediatric questions is statistically significant (p < 0.05). In all of these cases, performance was lower in the pediatric domain. The largest performance gap is seen with Gemini 2.0 Flash, with a drop of approximately 10.3 percentage points (81.24% vs 70.96%), followed by GPT 3.5-turbo-0125 with a decrease of about 9 percentage points (64.54% vs 55.59%). Even high-performing models like Grok 2 show a significant, though smaller, decline (around 2.6 percentage points, 89.18% vs 86.57%, p = 0.02). Conversely, three models—ChatGPT-4o, Gemini 1.5 Pro, and Claude 3.5 Sonnet (version 20241022)—do not exhibit a statistically significant difference in performance between the two medical domains. Particularly noteworthy is the performance of ChatGPT-4o, which achieves nearly identical accuracy scores (83.57% vs 83.18%[13], p = 0.80), indicating remarkable robustness and balance in handling both adult and pediatric content. For Gemini 1.5 Pro [8] and Claude 3.5 Sonnet [12], the observed differences (approximately 2.9 and 2.6 percentage points lower for pediatrics, respectively) do not reach the conventional threshold for statistical significance, with p-values of 0.07 and 0.06.

**Figure 1:**
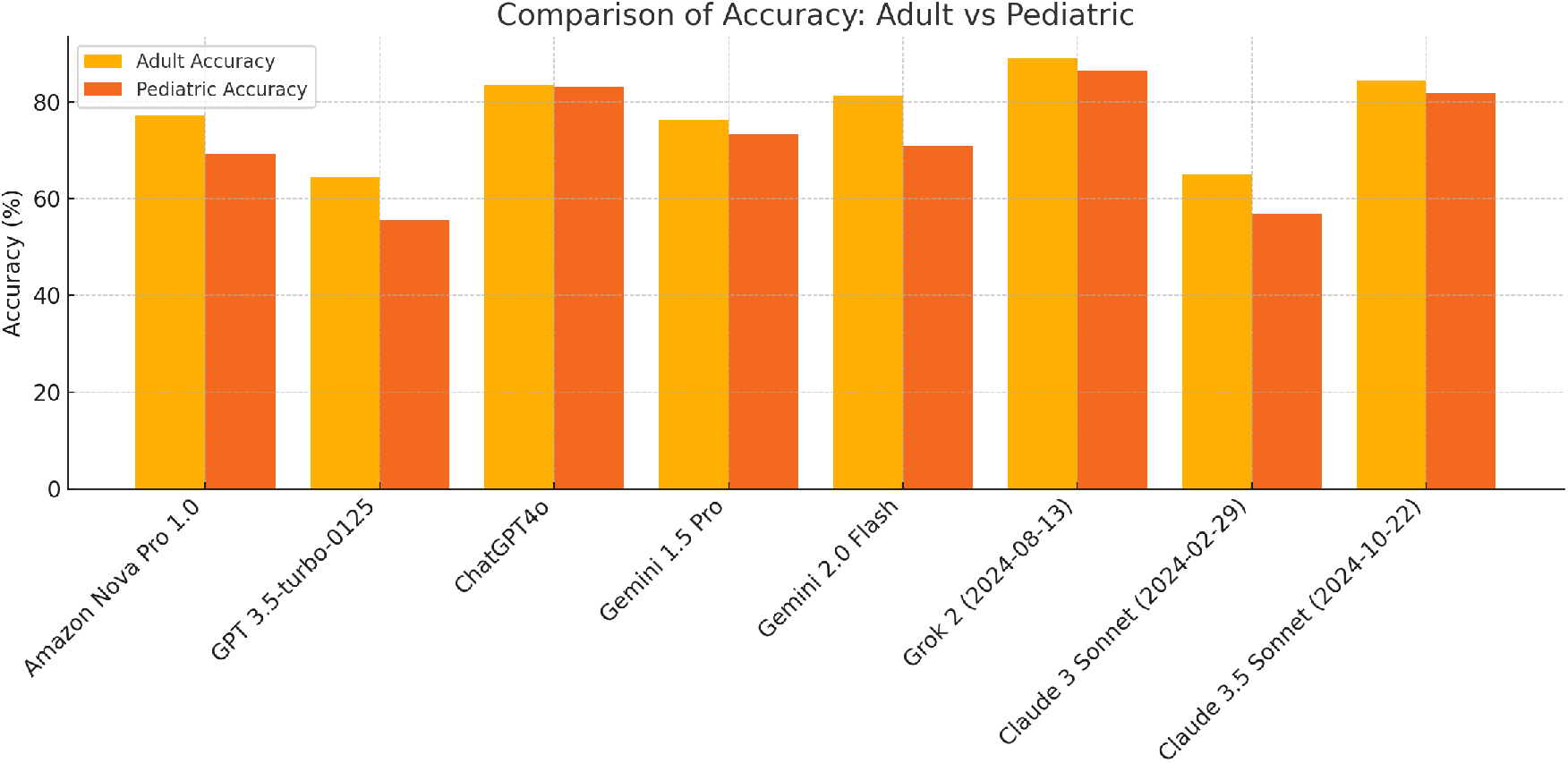
Bar chart comparing the performance of leading AI models in adult and pediatric contexts.

## 4 Discussion

The findings of this study offer a nuanced and multifaceted view of the initial hypothesis. On one hand, the fact that a majority of tested models (five out of eight) exhibit a statistically significant performance drop on pediatric questions partially supports our assumption. On the other hand, the absence of such a discrepancy in top-performing models like ChatGPT-4o and Claude 3.5 Sonnet, as well as Gemini 1.5 Pro, introduces layers of complexity that merit deeper exploration. The observed decline in models such as Amazon Nova Pro 1.0 [5], GPT 3.5-turbo-0125 [6], and the earlier version of Claude 3 Sonnet [11] may reflect limitations tied to factors such as model size (number of parameters), underlying architecture, or more likely, the composition of their pretraining data. It is plausible to hypothesize that the large text corpora used to train these models contained quantitatively or qualitatively less pediatric-specific medical content compared to adult medicine, making it more difficult for the models to accurately acquire and apply specialized knowledge like age-specific reference ranges or pediatric drug dosages. GPT 3.5-turbo, known to be less performant than its successors, might exemplify this limitation more prominently. What is particularly intriguing is that even powerful and recent models like Gemini 2.0 Flash [9] and Grok 2 [10] show a significant, albeit smaller in the case of Grok 2, discrepancy. This suggests that raw computational power or recency alone do not ensure uniform coverage across all specialized medical subdomains. It may be that pediatric knowledge, with its intrinsic peculiarities, presents a particularly difficult generalization challenge even for advanced architectures. Alternatively, training strategies may be optimized for average performance across a wide range of knowledge domains, leading to relative under-representation or reduced “depth” in the understanding of niche areas such as pediatrics. The substantial drop seen in Gemini 2.0 Flash (-10.3 points) is especially striking and may point to specific architectural or pretraining data features unique to this latency-optimized version, compared to others in the same family, such as Gemini 1.5 Pro. The latter employs a Mixture-of-Experts architecture [8], where the model is divided into numerous independent modules—so-called “experts”—and only a subset is activated for each input, through a dynamic gating network. This allows for a dramatic increase in total model capacity without a corresponding rise in computational load per inference, as each input activates only a few experts. In Gemini 1.5 Pro, this architecture supports higher efficiency in processing long contexts and increased accuracy on complex multimodal tasks.

On the other hand, the exceptionally balanced performance of ChatGPT-4o [7] (p = 0.80) is a remarkable outcome. This model appears to have achieved a level of training or architectural refinement that allows it to handle both domains with equal effectiveness. This could be attributed to an extremely large and diverse pretraining dataset that includes adequate representation of pediatric literature and practice, or to particularly effective fine-tuning techniques such as Reinforcement Learning from Human Feedback (RLHF), which promote generalization and factual accuracy even in specialized domains. The cases of Gemini 1.5 Pro and Claude 3.5 Sonnet [12] are equally enlightening. Both are recent models known for their large context windows—though not directly leveraged in simple MCQ tasks—and advanced capabilities. Their p-values of 0.07 and 0.06, while not formally significant, suggest a slight trend toward lower performance in pediatrics. One could speculate that these models represent a new generation progressively closing the gap, thanks to incremental improvements in training techniques, data quality and coverage, or architectural design. The lack of statistical significance may be due to sample power or may reflect a real but smaller gap compared to earlier models or other architectures such as Gemini 2.0 Flash. Claude 3.5 Sonnet, being the latest release from Anthropic at the time of testing, appears to improve upon its predecessor (Claude 3 Sonnet [11]) specifically in this aspect of differential performance. It is crucial to emphasize that no simple linear correlation emerges between a model’s release date or overall performance and the presence or absence of a significant domain-specific gap. The comparison between Gemini 2.0 Flash (with a gap) and Gemini 1.5 Pro (without one), or between Claude 3 Sonnet (with a gap) and Claude 3.5 Sonnet (without a significant gap), clearly illustrates this. This reinforces the notion that specific performance depends on a complex interplay of technical factors: model architecture (e.g., standard Transformer vs. Mixture-of-Experts), exact model size, detailed composition and curation of the pretraining dataset (especially domain representation), and fine-tuning and alignment strategies. It is worth noting that the deliberate choice to use each model’s default interface and hyperparameters was intentional. This decision aims to simulate a realistic use case, in which a clinical user—typically not an expert in LLMs or in parameters such as “temperature” or “Top P”—interacts with the tool “as is” via common platforms. Therefore, the results may more accurately reflect the performance a typical medical user, lacking expertise in prompt engineering or hyperparameter tuning, might expect in practical settings [14, 15]. While customized tuning could potentially improve performance, this approach ensures greater generalizability of the findings to standard use conditions. Nonetheless, it is essential to contextualize the interpretations proposed in this discussion. Our arguments regarding the impact of pretraining data (in terms of pediatric representation) and model characteristics (size, type, etc.) remain informed speculation. Given the closed-source nature of most of the models evaluated, we lack access to detailed technical specifications, pretraining datasets, or fine-tuning procedures. As such, we cannot empirically confirm whether, or to what extent, the presence or absence of adequate pediatric content in the training corpus or specific architectural choices truly contribute to the observed performance differences. Further research—potentially involving open-source models with transparent documentation or direct collaboration with developers of closed-source systems—would be necessary to validate these hypotheses.

### 4.1 Implications and Future Directions

These findings have significant practical implications for the adoption of LLMs in healthcare. They reaffirm the critical need for domain-specific evaluation prior to integrating such tools into clinical or educational workflows—especially in highly specialized areas like pediatrics. Relying on general performance metrics may obscure critical weaknesses in specific subdomains. The observed variability among models also underscores that not all LLMs are interchangeable for specialized medical tasks; model selection should be guided by evidence of performance within the intended use context. Future directions include:

- **Qualitative Error Analysis:** Examine pediatric questions where models failed to identify error patterns (e.g., dosage mistakes, age-specific reference ranges, rare syndromes) and understand the root causes of performance gaps.
- **Domain-Specific Fine-Tuning:** Assess the effectiveness of targeted fine-tuning techniques using pediatric-only text corpora to improve domain-specific performance.
- **Evaluation Across Diverse Tasks:** Extend the analysis beyond MCQs, using datasets that include complex clinical cases, open-ended questions, or tasks involving unstructured data interpretation to assess more advanced clinical reasoning abilities.
- **Ongoing Benchmarking:** Reassess model performance regularly as new LLMs become available, given the rapid pace of progress in the field.

## 5 Study Limitations

Some intrinsic limitations of this work must be acknowledged.

- **Single Dataset:** The use of a single dataset (MedQA), though widely adopted for benchmarking medical LLMs, may not capture the full complexity of medical reasoning or be representative of other evaluation types (e.g., open clinical cases, data interpretation, dialogic interaction). Results may not generalize to other datasets or clinical tasks.
- **Question Classification:** The a priori classification of questions into “Adult” and “Pediatric,” although based on objective criteria (patient age), may involve ambiguity or overlap in some cases, introducing minimal subjectivity. No formal inter-rater validation was performed.
- **Rapid LLM Evolution:** LLMs are rapidly evolving; the tested versions (with their specific knowledge cutoffs and release dates) may soon be superseded by newer models with different capabilities. Results reflect a specific temporal snapshot.
- **Nature of MCQs:** Evaluation through MCQs primarily measures factual recall and application of specific rules, but may not fully reflect complex clinical reasoning, differential diagnosis, uncertainty management, or treatment planning in real-world scenarios.
- **Prompting Strategy:** The specific prompt formulation used (“return only the letter…”) may have influenced responses. Different prompting strategies (e.g., few-shot learning, chain-of-thought) could yield different results, although consistency was maintained across all models.
- **Default Hyperparameters:** As discussed, the use of platform-default hyperparameters, while realistic, may not represent optimal performance achievable with customized configurations.

## 6 Conclusion

In conclusion, this comparative study highlighted significant heterogeneity in the performance of various LLMs when answering pediatric versus adult medical questions from the MedQA dataset. While some models, including recent and powerful ones, support the hypothesis of specific difficulty in the pediatric domain with a statistically significant drop in accuracy, others—particularly the most recent and advanced platforms like ChatGPT-4o—demonstrate a much more balanced capability across both domains. These findings suggest that while challenges in representing and reasoning about pediatric knowledge still exist for some LLMs, technological advancements are leading to the development of models with increasingly robust medical generalization. However, the observed variability among models, even within the same “family” (e.g., Gemini), calls for caution and highlights the crucial importance of rigorous, domain-specific validation before deploying any LLM in pediatric clinical or educational contexts. Future research should aim to better understand the roots of observed discrepancies and explore strategies to enhance the reliability of LLMs in specialized medical domains.

## Data Availability

All data produced in the present study are available upon reasonable request to the authors

## References

[1] Liu, Lei & Yang, Xiaoyan & Lei, Junchi & Liu, Xiaoyang & Shen, Yue & Zhang, Zhiqiang & Wei, Peng & Gu, Jinjie & Chu, Zhixuan & Qin, Zhan & Ren, Kui. (2024). A Survey on Medical Large Language Models: Technology, Application, Trustworthiness, and Future Directions. 10.48550/arXiv.2406.03712.

[2] Kaggle. MedQA-USMLE: Multiple choice question answering based on the United States Medical License Ex. 2025. https://www.kaggle.com/datasets/evidence/medqa-usmle. Accessed 12 March 2025.

[3] What Disease Does this Patient Have? A Large-scale Open Domain Question Answering Dataset from Medical Exams. arXiv. 2020. 10.48550/arXiv.2009.13081.

[4] LLM Large Language Model Directory https://docsbot.ai/models. Accessed 12 April 2025.

[5] Amazon Artificial General Intelligence. The amazon nova family of models: Technical report and model card. Amazon Technical Reports, 2024. https://www.amazon.science/publications/ the-amazon-nova-family-of-models-technical-report-and-model-card. Accessed 12 April 2025.

[6] GPT-3.5 Turbo - https://platform.openai.com/docs/models/gpt-3.5-turbo. Accessed 12 April 2025.

[7] GPT-4o System Card - https://cdn.openai.com/gpt-4o-system-card.pdf. Accessed 12 April 2025.

[8] Google Gemini Team, et al. (2024). Gemini 1.5: Unlocking multimodal understanding across millions of tokens of context. arXiv. 10.48550/arXiv.2403.05530.

[9] Introducing Gemini 2.0: our new AI model for the agentic era - https://blog.google/technology/ google-deepmind/google-gemini-ai-update-december-2024/#ceo-message. Accessed 12 April 2025.

[10] Grok-2 Beta Release - https://x.ai/news/grok-2. Accessed 12 April 2025.

[11] Anthropic. The claude 3 model family: Opus, sonnet, haiku, 2023. https://www-cdn.anthropic.com/de8ba9b01c9ab7cbabf5c33b80b7bbc618857627/Model_Card_Claude_3.pdf. Accessed 12 April 2025.

[12] Claude 3.5 Sonnet - https://www.anthropic.com/news/claude-3-5-sonnet. Accessed 12 April 2025.

[13] chat.openai.com - https://chat.openai.com. Accessed 12 April 2025.

[14] Google AI Studio - https://aistudio.google.com/. Accessed 12 April 2025.

[15] Chatbot Arena (formerly LMSYS): Free AI Chat to Compare & Test Best AI Chatbots - https://lmarena.ai/. Accessed 12 April 2025.

